# Estimating critical care capacity needs and gaps in Africa during the COVID-19 pandemic

**DOI:** 10.1101/2020.06.02.20120147

**Authors:** Jessica Craig, Erta Kalanxhi, Gilbert Osena, Isabel Frost

## Abstract

**Objective:** The purpose of this analysis was to describe national critical care capacity shortages for 52 African countries and to outline needs for each country to adequately respond to the COVID-19 pandemic.

**Methods:** A modified SECIR compartment model was used to estimate the number of severe COVID-19 cases at the peak of the outbreak. Projections of the number of hospital beds, ICU beds, and ventilators needed at outbreak peak were generated for four scenarios – if 30, 50, 70, or 100% of patients with severe COVID-19 symptoms seek health services—assuming that all people with severe infections would require hospitalization, that 4.72% would require ICU admission, and that 2.3% would require mechanical ventilation.

**Findings:** Across the 52 countries included in this analysis, the average number of severe COVID-19 cases projected at outbreak peak was 138 per 100,000 (SD: 9.6). Comparing current national capacities to estimated needs at outbreak peak, we found that 31of 50 countries (62%) do not have a sufficient number of hospital beds per 100,000 people if 100% of patients with severe infections seek out health services and assuming that all hospital beds are empty and available for use by patients with COVID-19. If only 30% of patients seek out health services then 10 of 50 countries (20%) do not have sufficient hospital bed capacity. The average number of ICU beds needed at outbreak peak across the 52 included countries ranged from 2 per 100,000 people (SD: 0.1) when 30% of people with severe COVID-19 infections access health services to 6.5 per 100,000 (SD: 0.5) assuming 100% of people seek out health services. Even if only 30% of severely infected patients seek health services at outbreak peak, then 34 of 48 countries (71%) do not have a sufficient number of ICU beds per 100,000 people to handle projected need. Only four countries (Cabo Verde, Egypt, Gabon, and South Africa) have a sufficient number of ventilators to meet projected national needs if 100% of severely infected individuals seek health services assuming all ventilators are functioning and available for COVID-19 patients, while 35 other countries require two or more additional ventilators per 100,000 people.

**Conclusion:** The majority of countries lack sufficient ICU bed and ventilator capacity to care for the projected number of patients with severe COVID-19 infections at outbreak peak even if only 30% of severely infected patients seek health services.

This analysis reveals there is an urgent need to allocate resources and increase critical care capacity in these countries.

## Introduction

The COVID-19 pandemic is rapidly emerging across the African continent with over 100,000 confirmed cases and over 2,600 deaths as of 01 June 2020 (World Health Organization). Overall disease prevalence has taken longer to accelerate in growth compared to other regions; however, most African countries are now reporting community transmission, and several countries have started to see week-on-week exponential increases in confirmed cases (Dong). Estimates of COVID-19 incidence and mortality rates are complicated by low testing capacity in many countries, and it has been suggested that COVID-19 deaths are being under-reported in some areas (Kavanagh; Maclean). Previous assessments of health systems and critical care capacity, including a COVID-19 Readiness Survey conducted by the World Health Organization in March 2020, suggest many African countries are not adequately equipped to cope with surges in demand for hospital beds, intensive care unit (ICU) beds, and mechanical ventilators (World Health Organization).

Critical care capacity, needs, and overall COVID-19 pandemic readiness in Africa remain unclear. A more complete understanding of COVID-19 and local case burden patterns is also needed, given that most countries are at the beginning of their respective outbreaks. Measures taken by other countries to mitigate COVID-19 transmission, such as curfews and lockdowns, may not be feasible or effective across Africa given various social, economic, political, and demographic factors such as high population density in urban cities, slums, and refugee camps; high proportion of the workforce employed in the informal sector, and high rates of poverty (Partnership for Evidence-Based Response to COVID-19; Hopman; International Labour Organization). As a result, it has been predicted that the COVID-19 case burden may, at present and in the long term, be higher across Africa than in high-income countries (Wells; World Health Organization).

Assessing current national health system capacities and future needs in the COVID-19 pandemic context is challenging given the lack of publicly available data. This is further complicated by uncertainties and differences in care-seeking behaviour across the continent. In some cases, even where hospital facilities are available, they may be underutilised due to affordability or safety concerns (Nwankwo; Wambui). Despite uncertainties around disease transmission and challenges in assessing health systems needs, estimates are essential for public health planners and policymakers to make evidence-based decisions on the distribution of resources in the coming months as COVID-19 progresses across Africa.

In a previous study, we compiled a database describing various aspects of critical care capacity in Africa as relevant to the COVID-19 pandemic (Craig). Separately, we utilized a previously described SECIR compartment model to estimate cumulative and peak case burden in all African countries (Center for Disease Dynamics, Economics, & Policy). In this paper, we combine these analyses and use the best available data on current critical care capacity for 52 African countries and projections for peak case burden to describe national capacity shortages and outline needs for each country to adequately respond to the pandemic.

## Methods

### Estimates of Current Critical Care Capacity

Data on essential components of critical care capacity relevant to COVID-19 treatment, such as number of hospital beds, ICU beds, and ventilators were compiled from various sources including published governmental and non-governmental reports, scientific literature, local and international media, and in-country informants including government or public health officials and other local researchers and healthcare workers, as previously described (Craig).

### COVID-19 Case Projections

A modified Susceptible – Exposed – Contagious – Infected – Recovered (SECIR) compartment model was used to predict national COVID-19 case burden for African countries based on current case data and lockdown interventions (Dong).

The model’s parameters and assumptions have been previously described (Center for Disease Dynamics, Economics, & Policy). Briefly, model parameters include a basic reproductive number of 2.74, an incubation period of three days, and a six percent rate of progression to severe disease. In addition, the model assumes an asymptomatic clearance period of 3 days and a symptomatic clearance period of 5 days. Duration and timing of lockdown interventions by country were obtained from a review of local and international news media, and country and diplomatic mission reports. Peak estimates were based on the assumption that lockdowns decreased COVID-19 transmission by 25%, and, after lockdown was lifted, disease transmission returned to 90% of the pre-lockdown value. The parameters used to estimate severe cases in Africa were based on hospitalisation rates in regions where most data on COVID-19 epidemiology was available (Lin; Liu; Bi; J. L. Wu). The impact of age and comorbidities are not explicitly included in the model but are contained as a function of the estimated number of people who will have severe disease.

### Estimating Critical Care Capacity Needs and Gaps in Current Capacity

We estimated the number of hospital beds, ICU beds, and ventilators needed during the peak of each country’s respective COVID-19 outbreaks assuming changes to disease transmission under the lockdown scenario described above.

It was assumed that all people with severe COVID-19 infections would require hospitalization, that 4.72% of severe cases would require ICU admission, and that 2.3% of severe cases would require mechanical ventilation (Guan; European Centre for Disease Prevention and Control; Emami; Team, Severe Outcomes Among Patients with Coronavirus Disease 2019 (COVID-19) – United States, February 12-March 16, 2020). The number of cases in need of hospital beds was derived by subtracting the number of cases in need of ICU beds from the total number of cases assumed to require hospitalization.

Given that access to and utilization of health services may be limited across the continent and that patients may opt for home treatment versus inpatient care, we generated estimates of hospital bed, ICU bed, and ventilator needs and gaps for four scenarios assuming that 30, 50, 70, or 100% of patients with severe COVID-19 symptoms will seek health services (Akpunne; Naanyu; Aantjes; Hopkins).

Comoros and Lesotho were excluded from all analyses due to lack of COVID-19 case data. Dependent territories in the Africa region were also excluded.

Due to lack of data on current national capacities, gaps in hospital bed capacity at outbreak peak were not calculated for two countries (Eswatini and South Sudan), for ICU bed capacity for four countries (Benin, Equatorial Guinea, Madagascar, and Mozambique), and gaps in ventilator capacity were not calculated for six countries (Benin, Republic of the Congo, Eritrea, Malawi, Mauritius, and the Seychelles). However, hospital bed, ICU bed, and ventilator needs at outbreak peak were still estimated for these countries.

## Results

### COVID-19 Case Projections

Across the 52 countries included in this analysis, the average number of severe COVID-19 cases projected at outbreak peak was 138 per 100,000 (Standard Deviation [SD]: 9.6) and ranged from 102 per 100,000 in Sao Tome and Principe and Egypt to 145 per 100,000 in Equatorial Guinea and Uganda (Appendix 1).

### Hospital Bed Capacity

The average number of hospital beds needed at the peak of respective national COVID-19 outbreaks across 52 countries assuming 100% of infected patients with severe symptoms seek out health services was 131.7 beds per 100,000 people (SD: 9.2) ranging from 96.8 per 100,000 in Egypt to 137.8 beds per 100,000 in Equatorial Guinea. The average number of hospital beds needed at the peak of the outbreak was 39.5 (SD: 2.8), 65.9 (SD: 4.6), and 92.2 (SD: 6.4) per 100,000 people assuming only 30, 50, and 70% of infected patients seek health services, respectively. Hospital bed needs and gaps at the peak of respective national COVID-19 outbreaks are fully described in Appendix 2.

Comparing current national capacities to estimated needs at outbreak peak, we found that 31 of 50 countries (62%) do not have a sufficient number of hospital beds per 100,000 people if 100% of patients with severe infections seek out health services; if only 30% of patients seek out health services then 10 of 50 countries (20%) do not have sufficient capacity (Figure 1). (These estimates assume that all hospital beds are empty and are available for use by patients with COVID-19.)

**Figure 1.**
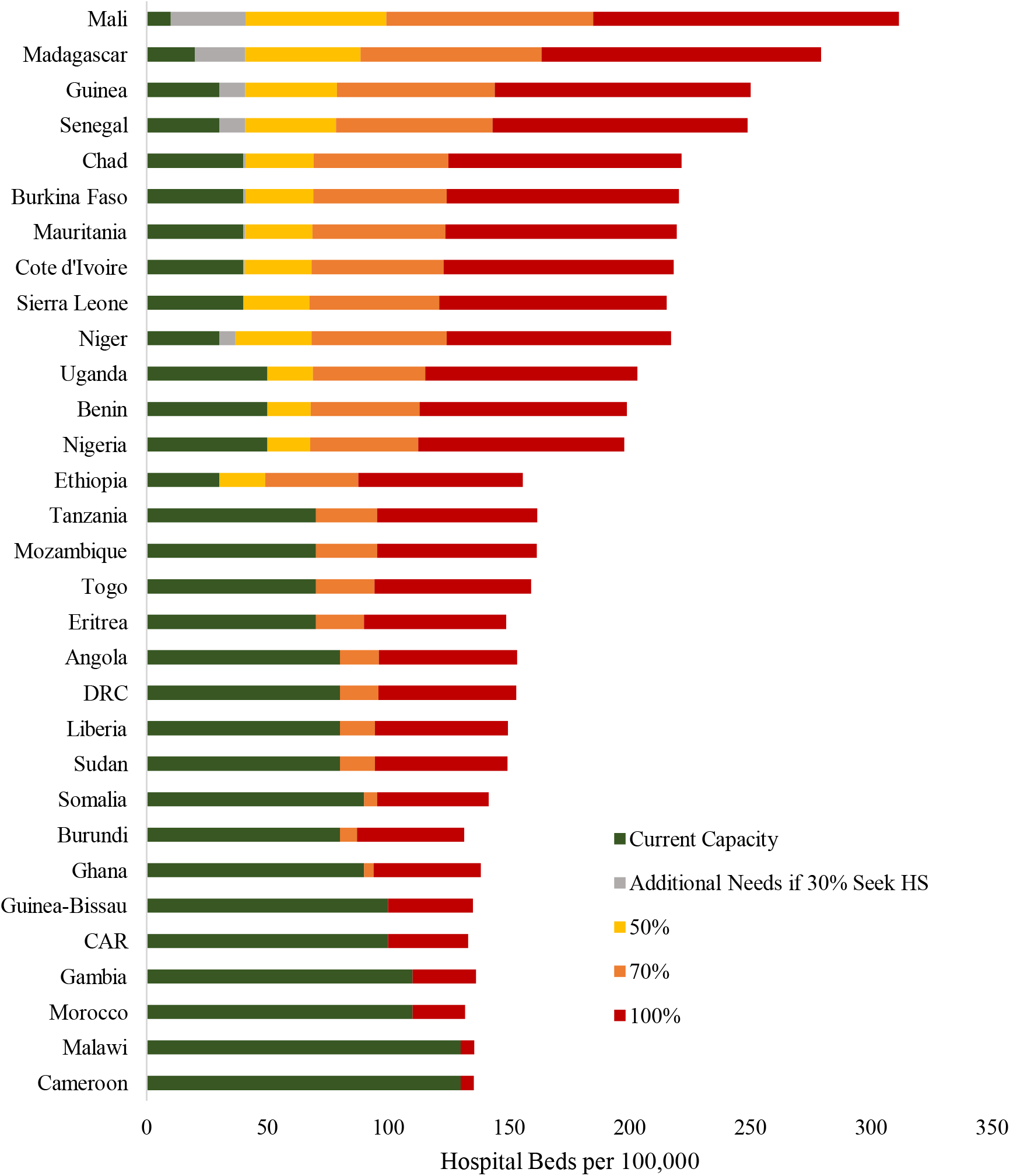
Hospital Beds Required per 100,000 at Outbreak Peak for 31 Countries with Current Capacity Gaps

Mali, Madagascar, Senegal, Niger, and Guinea have the largest gap between current hospital bed capacity and predicted need during outbreak peak under the scenario when only 30% of people with severe COVID-19 infections seek out health services; our findings suggest that Senegal, Niger, and Guinea required more than 1,000 additional hospital beds and Mali and Madagascar more than 5,000 additional hospital beds. If 100% of people with severe infections seek health services at outbreak peak, then each of those countries would require over 10,000 additional hospital beds.

### ICU Bed Capacity

The average number of ICU beds needed at outbreak peak across the 52 included countries ranged from 2 per 100,000 people (SD: 0.1) when 30% of people with severe infections access health services to 6.5 per 100,000 (SD: 0.5) assuming 100% of people seek out health services. ICU bed need at peak was highest in Equatorial Guinea and Uganda and lowest in Egypt and Sao Tome and Principe.

If 30% of severely infected patients seek health services at outbreak peak, then 34 of 48 countries (71%) do not have a sufficient number of ICU beds per 100,000 people to handle projected need (Figure 2). If 100% of patients with severe COVID-19 infections seek health services, only 6 countries (Botswana, Egypt, Cameroon, Gabon, Mauritius, and Seychelles) have sufficient ICU bed capacity per 100,000 to care for COVID-19 patients assuming all ICU beds are available while 32 other countries (67%) need more than 5 additional ICU beds per 100,000 people to adequately care for severely infected COVID-19 patients at the peak of the outbreak. Predicted needs and gaps in ICU bed capacity at the peak of respective national COVID-19 outbreaks are fully described in Appendix 3.

**Figure 2.**
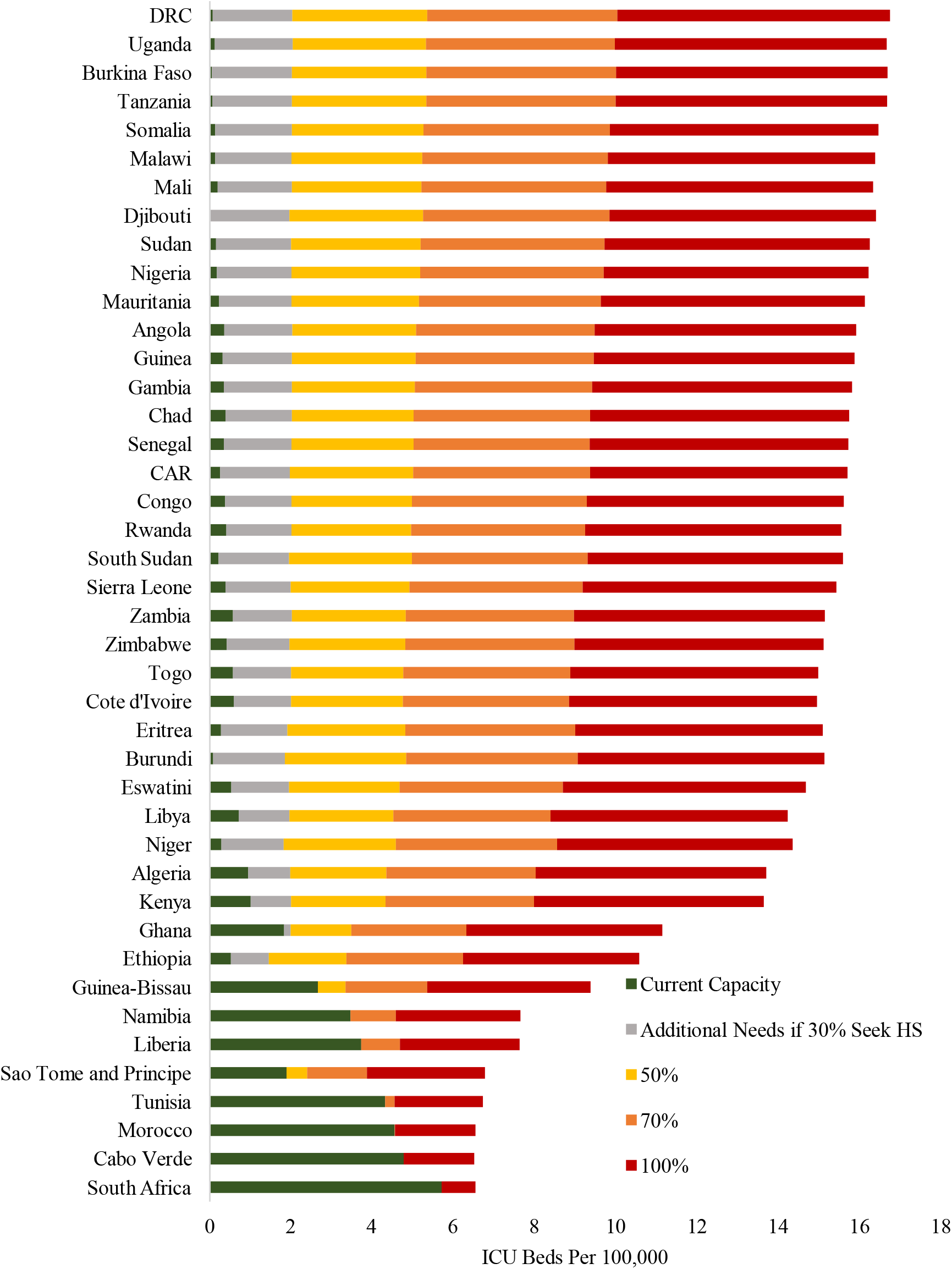
ICU Beds Required per 100,000 at Outbreak Peak for 42 Countries with Current Capacity Gaps

In order to meet the projected ICU bed needs at outbreak peak if 100% of severely infected patients seek health services, our findings suggest DRC and Ethiopia require approximately 5,000 additional ICU beds while Nigeria would need over 12,000 additional beds.

### Ventilator Capacity

Across the 52 countries, the average number of ventilators needed when there is a peak number of severe infections ranged from 0.95 per 100,000 (SD: 0.1) assuming 30% of severely infected people seek health services to 3.18 per 100,000 (SD: 0.2) assuming 100% of severely infected people seek health services. Projected needs and gaps in current ventilator capacity are fully described in Appendix 4.

If 100% of severely infected individuals seek health services, then only four countries (Cabo Verde, Egypt, Gabon, and South Africa) have a sufficient number of ventilators to meet national needs, assuming all ventilators are functioning and available for COVID-19 patients, while 35 other countries require two or more additional ventilators per 100,000 people. Assuming only 30% of people with severe COVID-19 infections seek health services, then 35 of 46 (76%) still lack sufficient ventilator capacity to respond to projected need at outbreak peak. Gaps in current capacity and projected need for the other 42 countries are summarized in Figure 3.

**Figure 3.**
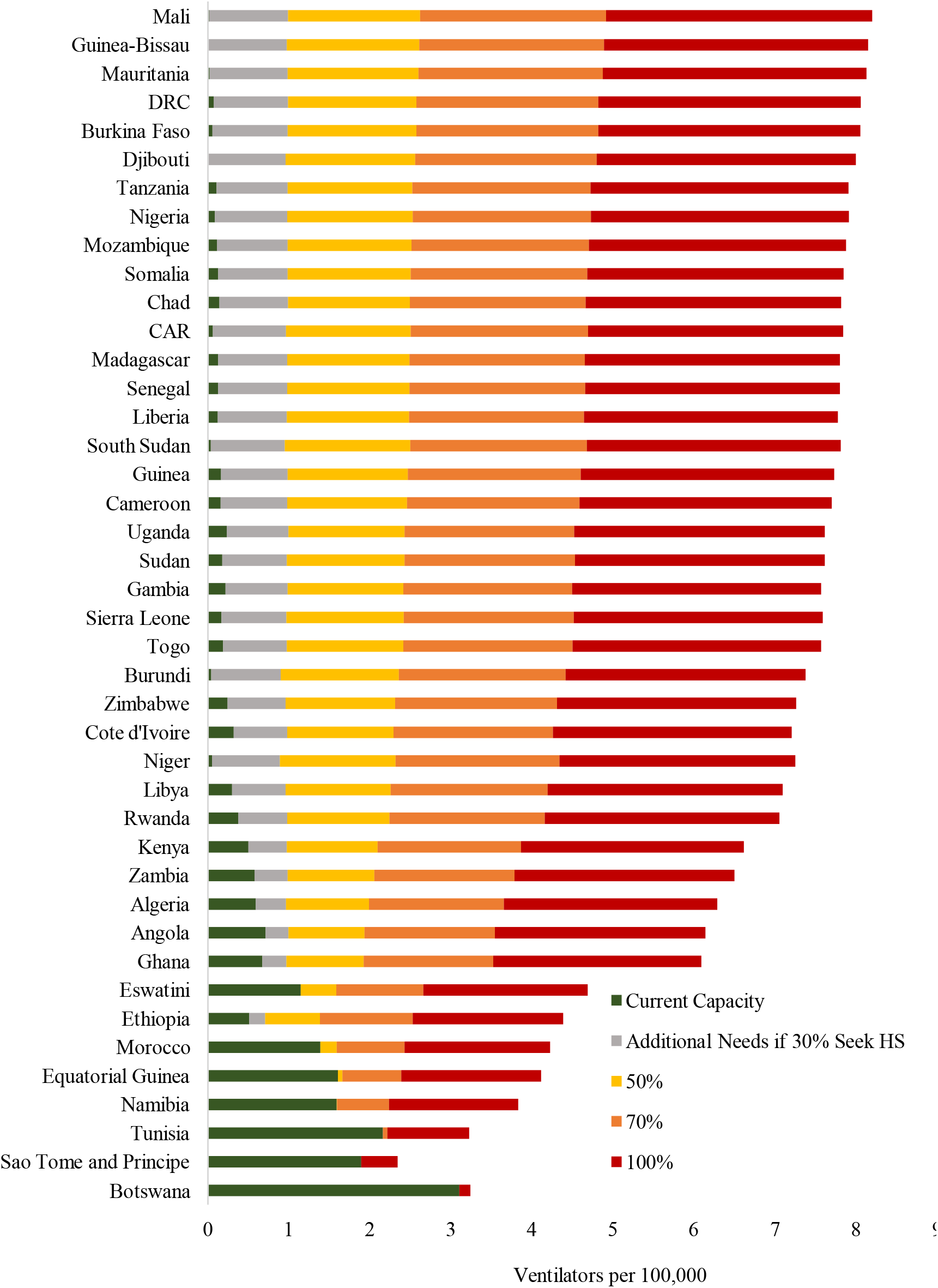
Ventilators Required per 100,000 at Outbreak Peak for 42 Countries with Current Capacity Gaps

Guinea-Bissau, Mali, and Mauritania have the highest gaps when comparing the current number of ventilators per 100,000 people to projected peak need under all health access scenarios.

## Discussion

To date, this is the most comprehensive assessment of critical care gaps and needs across African countries under COVID-19 pandemic conditions. However, this analysis has several limitations. First, national critical care estimates may, in some instances, over- or under-represent the true critical care capacity of a country (i.e. estimates from government published reports may only reflect critical care capacity at public sector health facilities designated to care for COVID-19 patients, excluding those from the private sector). In addition, since information on critical care components were collected from different sources and represented estimates from different time points, the number of hospital beds may include a proportion of available ICU beds. Peak estimates are generated from an SECIR model which assumes populations are well-mixed and this can lead to overestimation of the peak need.

We assume that all hospital beds, ICU beds, and ventilators are functioning and available exclusively for people with COVID-19 infections, an assumption which is unlikely to reflect reality. Anecdotally, healthcare workers from multiple African countries report that health facilities are seeing significantly fewer patients compared to pre-pandemic months as patients face increased challenges accessing health services or fear becoming infected with the virus at a health facility.

Data on hospital bed, ICU bed, and ventilator distribution were unavailable, and this analysis assumes that they are evenly distributed across populations; in reality, healthcare infrastructure is typically clustered in capital cities and other urban centers (Okafor; Hoeven; Oloyede; World Health Organization). In addition, the number of hospital beds, ICU beds, and ventilators may not reflect the quality of healthcare; however, these are useful indicators for gauging the capacity of a healthcare system.

Model assumptions and variations in testing rates across the different countries may partially explain some of the differences between projected and reported cases. Estimation of disease severity across Africa comes with the challenges of accounting for multiple factors that could have negative or positive effects in the progression of the outbreak. For example, a relatively young population structure could mean that the percentage of severe COVID-19 cases will be lower than that estimated from data from China, Europe, and the US as old age has been shown to be an important risk factor for both disease severity and mortality (CDC COVID-19 Response Team; J. L. Wu; Cohen). However, despite having a younger population, factors such as high incidence of immunocompromising diseases, malnourishment and limited access to healthcare may increase the vulnerability of the population towards COVID-19.

Needs and gaps in current critical care capacity described here consider only the projected burden from those with severe COVID-19 infections. However, the pandemic and subsequent measures to mitigate disease transmission will likely impact all aspects of national health systems; for instance by disrupting bednet campaigns against malaria, reducing access to antiretroviral therapies (ART), and stifling essential supply chains thereby potentially increasing malnutrition rates (WHO Africa). It is important to consider that the pandemic is likely to further increase demand for non-COVID-19 health services and will further exacerbate existing gaps in health systems capacities (Hogan; Anthem).

Despite the limitations related to these estimates, the overall analysis reveals a lack of coping capacity in the face of the COVID-19 pandemic for many African countries and highlights the immediate need for investment in critical care infrastructure. Although the disease seems to be progressing at a slower rate across the African continent, it is predicted to persevere in the coming months, and therefore, rapid improvement of critical care capacity could assist in the treatment of those with severe infections, not only during the current pandemic wave but in the likely second and third waves (World Health Organization Africa). Furthermore, for countries where national critical care capacities may be adequate, ensuring equitable access across demographies, geographical locations, and socioeconomic classes is imperative.

## Data Availability

All data is or has been made publicly available.

